# Monitoring the Covid-19 epidemics in Italy from mortality data

**DOI:** 10.1101/2020.05.07.20092775

**Authors:** Daniele del Re, Paolo Meridiani

## Abstract

The mortality data can be used as an alternative source to monitor the status of Covid-19. We have studied a dataset including deaths up to the fourth week of April. There is a large excess, more pronounced at the beginning of the pandemic, showing a difference in age and gender compared to the Covid-19-confirmed cases. The study indicates that mortality information can be used to provide a less biased time profile of the pandemic.

---

The monitoring of the coronavirus pandemic (Covid-19) is usually based on indicators, such as the number of new positive cases per day, which heavily depend on the number of tests administered to the population. Since the number of tests increases as the disease spreads, the resulting epidemic curves are unable to fully represent the actual evolution of the Covid-19 pandemic in a given region [1]. In particular, in Italy the number of tests made has increased by more than a factor 4 during March 2020, as a combined result of changing the guidelines and the capacity to perform them. All subsequent epidemiological indicators, number of infected, recovered and dead, have been all affected by biases which are difficult to be estimated and corrected in absence of further information about the tests.

Overall mortality data can be used instead as an alternative source to monitor the status of Covid-19. The potential of mortality data is well known and discussed in several papers [2,3,4,5]. The dataset under study is provided by Istituto Nazionale di Statistica (ISTAT) [6] and Sistema di Sorveglianza Mortalità Giornaliera (SiSMG) [7]. It provides the number of deaths per day divided in age categories and gender, up to 4th April 2020, for ISTAT data, and up to 25th April for SiSMG data, i.e. after about two months from the first official direct transmission Covid-19 case reported in Italy. The dataset also contains the historical data, for the same months, for the years 2015-2019. A selection is applied to the municipalities included in the ISTAT dataset: only towns presenting an excess of 20% with respect to the historical average are present, inducing a potential bias which decreases with the dataset coverage. The combined dataset (ISTAT+SiSMG) refers to about 50% of the infected population, taking into account the regional spread of the epidemics. Big cities, like Milan, Turin and Rome are present. Lombardy, which accounts for about one half of the Covid19 deaths in Italy, has the largest coverage, around 72%. Mortality data are also being made available in other countries [8]: these data are not publicly available in a form that allows a complete statistical analysis.

The time-series data for the mortality excess, for different cities and regions, have been obtained by subtracting an average historical model, taking into account the seasonal mortality trend. This is obtained from the average of 2015-2019, normalised to the number of deaths observed in the first 3 weeks of February. Such a model is designed to best extrapolate the observed mortality rate at the beginning of February 2020 into March and April. As an example, the time-series before (left) and after (right) the historical model subtraction is shown in Fig. 1 for the city of Brescia. The same procedure can be applied to any data category, in particular to other cities and regions. The mortality excess is compared with the official Covid-19 deaths provided daily by the Italian *Protezione Civile* [9], as shown in the same figure (right). Confirmed deaths for Covid-19 are only available at regional granularity: values for cities have been obtained from the corresponding region, taking into account the fraction of active Covid-19 cases in the province as a function of time, and then rescaling for the population. This calculation assumes a uniform distribution of deaths within the same province; an uncertainty of 20% is assigned, obtained from a comparison between this estimate and the real value for a few cities where mortality data was available.

**Fig. 1:**
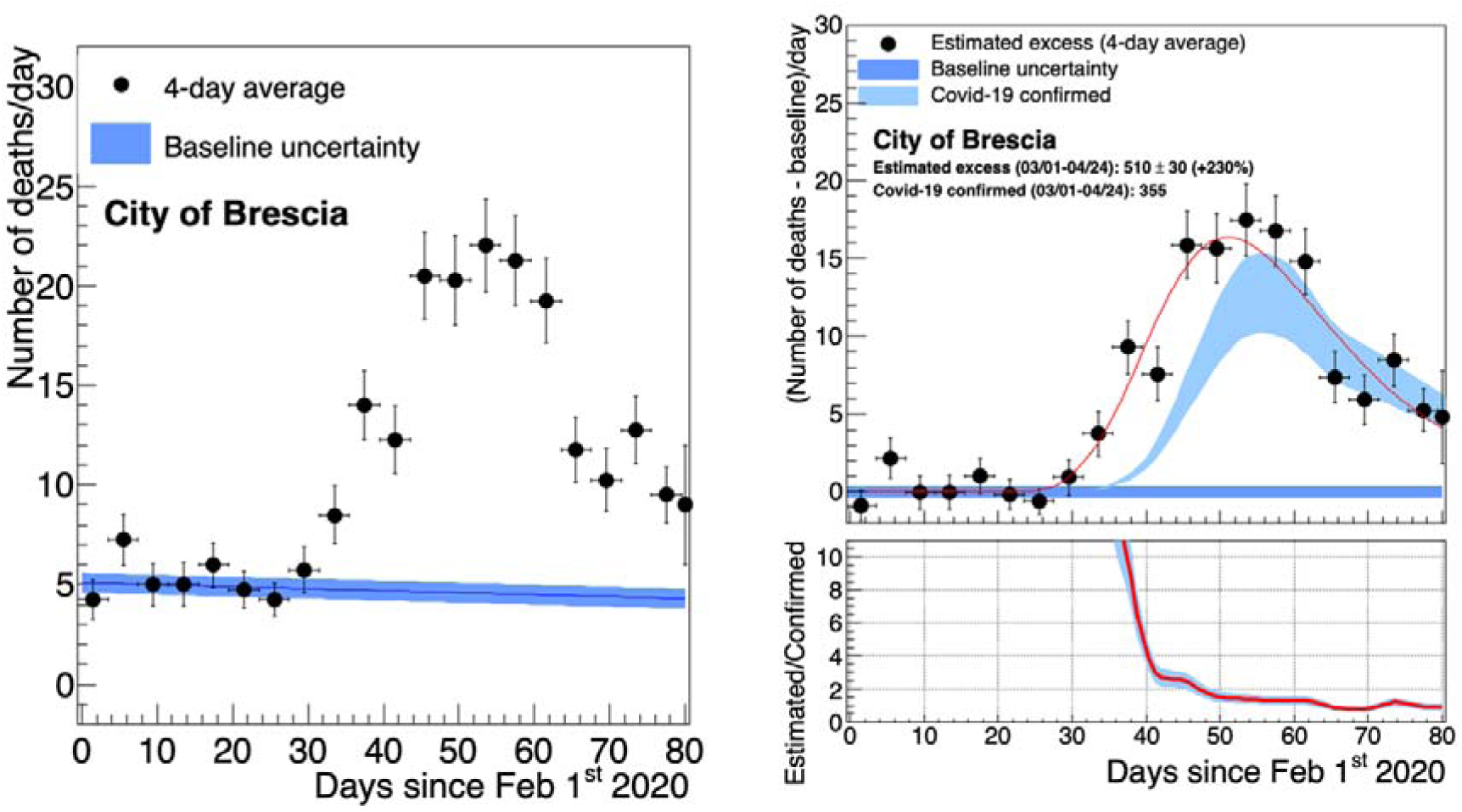
Left. distribution of deaths in Brescia as a function of time. Right. baseline-subtracted distribution of deaths in Brescia compared to the official Covid-19 distribution. The ratio of the two distributions is displayed below.

**Fig. 2:**
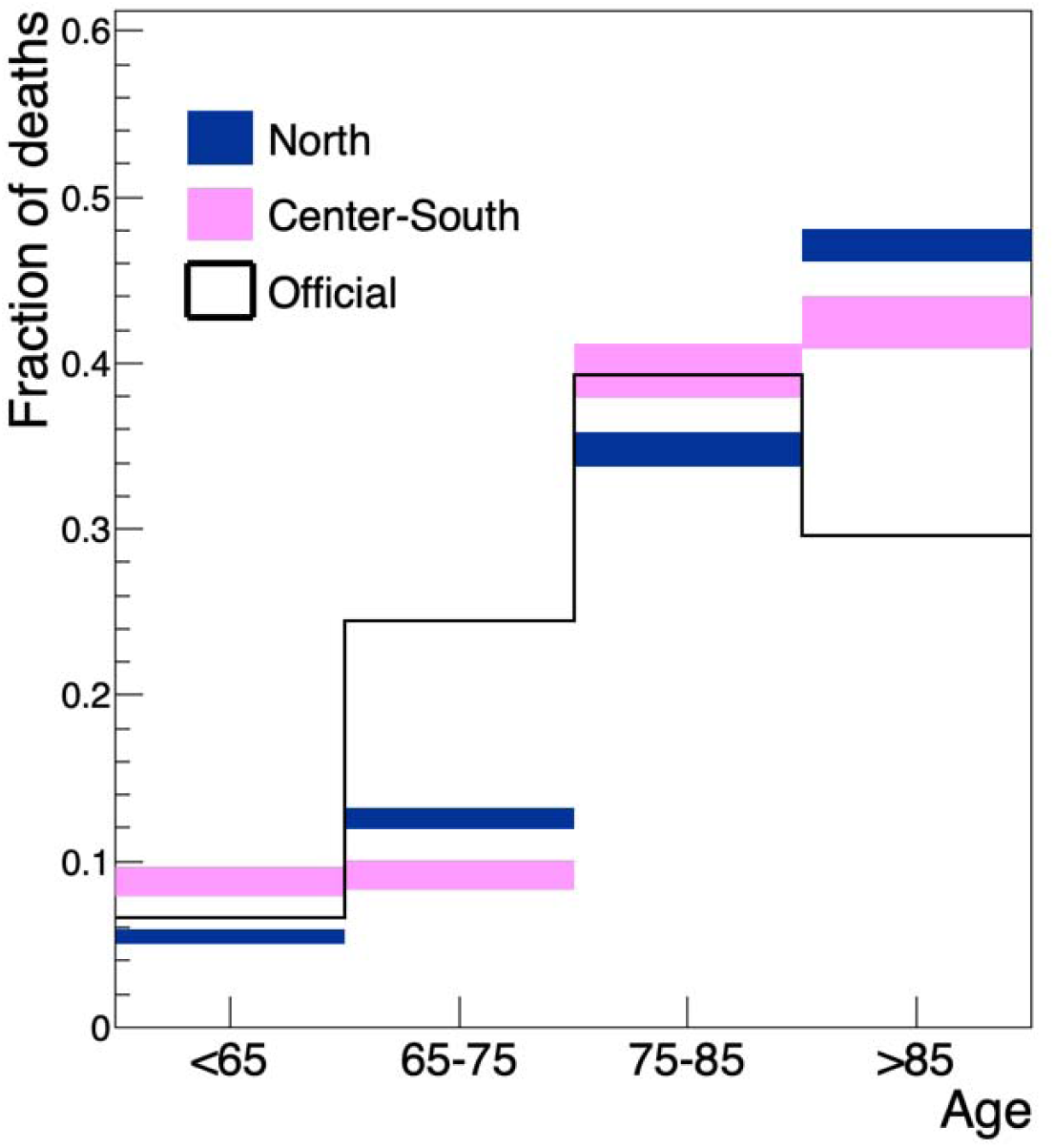
Baseline-subtracted distribution of the age at death compared to the official Covid-19 distribution.

The mortality excess time-series seems to follow the laboratory-confirmed cases, with the start of the raise at the end of February, when the first Italian case of Covid-19 has been reported. The excess of mortality is higher than the confirmed Covid-19 cases at early days of the pandemic, indicating that at the beginning of the outbreak a large fraction of cases hasn’t been identified by the official monitoring. Later, the two values tend to be closer, suggesting that the effective sampling of tested people has increased with time. Similar behaviour is observed for other cities and regions. A large spread of the ratio between the overall mortality excess and confirmed Covid-19 deaths is present: it is above two for many cities and, in some cases, turns out to be very large, as for the city of Genoa.

Tab. 1 summarizes these values. We also report the aggregated result for the full Italian sample.

**Tab. 1:** Excesses of mortality for several Italian cities compared to the official data.

| City (update 04/24/2020) | Mortality excess | Confirmed Covid-19 deaths | Population | Mortality excess pro-capite (‰) | Official Covid-19 mortality pro-capite (‰) |
| --- | --- | --- | --- | --- | --- |
| Milan | $2000 \pm 70$ | $900 \pm 180$ | 1352k | $1.48 \pm 0.05$ | $0.67 \pm 0.13$ |
| Brescia | $510 \pm 30$ | $350 \pm 70$ | 197k | $2.59 \pm 0.15$ | $1.78 \pm 0.36$ |
| Genoa | $1090 \pm 50$ | $300 \pm 60$ | 584k | $1.87 \pm 0.09$ | $0.51 \pm 0.10$ |
| Bologna | $320 \pm 40$ | $140 \pm 28$ | 388k | $0.82 \pm 0.10$ | $0.36 \pm 0.07$ |
| **North (sum)** | **$3920 \pm 110$** | **$1690 \pm 200$** | **2239k** | **$1.75 \pm 0.05$** | **$0.75 \pm 0.09$** |
| Perugia | $20 \pm 20$ | $10 \pm 2$ | 167k | $0.12 \pm 0.12$ | $0.06 \pm 0.01$ |
| Rome | $580 \pm 80$ | $180 \pm 36$ | 2873k | $0.20 \pm 0.03$ | $0.06 \pm 0.01$ |
| Bari | $150 \pm 30$ | $26 \pm 5$ | 3241k | $0.46 \pm 0.09$ | $0.08 \pm 0.01$ |
| Potenza | $38 \pm 15$ | $3 \pm 1$ | 672k | $0.57 \pm 0.22$ | $0.04 \pm 0.01$ |
| Palermo | $110 \pm 40$ | $18 \pm 4$ | 674k | $0.16 \pm 0.06$ | $0.03 \pm 0.01$ |
| Messina | $66 \pm 26$ | $13 \pm 4$ | 237k | $0.28 \pm 0.11$ | $0.05 \pm 0.01$ |
| **Center-South (sum)** | **$960 \pm 100$** | **$250 \pm 40$** | **4342k** | **$0.22 \pm 0.02$** | **$0.06 \pm 0.01$** |
| **Italy (ISTAT+SiSMG up to 04/04/2020)** | **$23400 \pm 500$** | **[7500,11400]** | **19304k** | **$1.21 \pm 0.03$** | **[0.38,0.59]** |

| City (update 04/24/2020) | Mortality excess | Confirmed Covid-19 deaths | Population | Mortality excess pro-capite (‰) | Official Covid-19 mortality pro-capite (‰) |
| --- | --- | --- | --- | --- | --- |
| Milan | 2000±70 | 900±180 | 1352k | 1.48±0.05 | 0.67±0.13 |
| Brescia | 510±30 | 350±70 | 197k | 2.59±0.15 | 1.78±0.36 |
| Genoa | 1090±50 | 300±60 | 584k | 1.87±0.09 | 0.51±0.10 |
| Bologna | 320±40 | 140±28 | 388k | 0.82±0.10 | 0.36±0.07 |
| **North (sum)** | **3920±110** | **1690±200** | **2239k** | **1.75±0.05** | **0.75±0.09** |
| Perugia | 20±20 | 10±2 | 167k | 0.12±0.12 | 0.06±0.01 |
| Rome | 580±80 | 180±36 | 2873k | 0.20±0.03 | 0.06±0.01 |
| Bari | 150±30 | 26±5 | 3241k | 0.46±0.09 | 0.08±0.01 |
| Potenza | 38±15 | 3±1 | 672k | 0.57±0.22 | 0.04±0.01 |
| Palermo | 110±40 | 18±4 | 674k | 0.16±0.06 | 0.03±0.01 |
| Messina | 66±26 | 13±4 | 237k | 0.28±0.11 | 0.05±0.01 |
| **Center-South (sum)** | **960±100** | **250±40** | **4342k** | **0.22±0.02** | **0.06±0.01** |
| Italy (ISTAT+SiSMG up to 04/04/2020) | 23400±500 | [7500,11400] | 19304k | 1.21±0.03 | [0.38,0.59] |

The effective coverage of the ISTAT+SiSMG dataset is taken into account to estimate the total number of official deaths. The uncertainty assigned reflects the partial coverage of the data.

Age and gender of the mortality excess differ significantly from the laboratory-confirmed Covid-19 deaths. The following Fig.2 shows the comparison between the distributions subtracted of historical data for cities in the Northern part of Italy (Brescia, Milan, Genoa, and Bologna), where the impact of the virus has been stronger with a mortality of about 0.2%, and for cities of Center-South (Rome, Perugia, Bari, Potenza, Palermo, Messina), where the mortality is about a factor 10 less, with the official Covid-19 deaths. Both distributions look quite different from the official data and also between themselves. This is an indication that an important fraction of the excess in deaths is due to older people, dying at home or at the hospice, without even accessing the healthcare system. The difference between North and Center-South may also suggest that in cities with a smaller Covid-19 mortality the impact of the deaths not directly due to the virus but connected to *lockdown* could be much more significant.

These data indicate that mortality information can be used to provide a less biased time profile of the pandemic. In general, there is a large excess of deaths, more pronounced at the beginning of the pandemic. The additional information regarding age, gender, and regional differences with respect to the official data may also provide a powerful handle to disentangle the actual Covid-19-related contribution from the deaths due to the stress of the healthcare system or to the reduced access to emergency rooms. The impact of this contribution should be minor, given that the difference between the excess of mortality and the official Covid-19 deaths becomes smaller, as time passes. If made available, the information about the cause of the death and the type of hospitalization, which are usually stored in a mortality database, will disentangle the different components, helping to identify the pure Covid-19 contribution. Similar comparisons can be also done between different countries, shedding light on the mechanisms of the spread of the infection and on its real impact.

## Data Availability

All results are obtained by using data by Istituto Nazionale di Statistica (ISTAT) and Sistema di Sorveglianza Mortalità Giornaliera (SiSMG)

https://www.istat.it/it/archivio/240401

http://www.deplazio.net/images/stories/SISMG/SISMG_COVID19.pdf

## References

[1] Monitoring the COVID-19 epidemic in the context of widespread local transmission, A. L. García-Basteiro, C. Chaccour, C. Guinovart, et al., The Lancet, https://doi.org/10.1016/S2213-2600(20)30162-4

[2] Monitoring excess mortality for public health action: potential for a future European network, A. Mazick et al., Eurosurveillance, Volume 12, Issue 1, 04 January 2007

[3] European all-cause excess and influenza-attributable mortality in the 2017/18 season: should the burden of influenza B be reconsidered?, J. Nielsen, L.S. Vestergaard, L. Richter, D. Schmid, NCBI, Clin Microbiol Infect. 2019 Oct;25(10):1266–1276

[4] Influenza-associated mortality determined from all-cause mortality, Denmark 2010/11–2016/17: The FluMOMO model, J. Nielsen, T.G. Krause, K. Mølbak, NCBI, Influenza Other Respir Viruses. 2018 Sep;12(5):591-604

[5] Excess mortality among the elderly in European countries, December 2014 to February 2015, Eurosurveillance, K. Mølbak, L. Espenhain, J. Nielsen, et al., Volume 20, Issue 11, 19 March 2015

[6] Dati di mortalità: cosa produce l’Istat, https://www.istat.it/it/archivio/240401

[7] Sintesi attività sistema di sorveglianza mortalità giornaliera per monitoraggio impatto Covid-19. Rapporto 1 Febbraio - 18 Aprile 2020, M.. Davoli, F. de’ Donato, M. De Sario, et al., EP Rev.

[8] The EuroMOMO network, https://www.euromomo.eu/

[9] https://github.com/pcm-dpc/C0VID-19

